# Systematic Review of Brain Mechanisms of Oral Multisensory Processing related to Oral Health

**DOI:** 10.1101/2024.08.10.24311798

**Authors:** Chia-Shu Lin

## Abstract

Oral functions related to eating, including mastication, swallowing, and taste, are fundamentally a multisensory experience, that relies on the crossmodal interaction of touch, gustation, temperature, pain, and proprioception. While a majority of studies focus on the multisensory processing related to speech, the brain mechanisms of oral multisensory processing related to eating have remained unknown. The current systematic review will summarize the findings from neuroimaging studies (mainly functional magnetic resonance imaging) focusing on the interaction of multiple sensory stimuli in human participants. Neuroimaging studies of human adults on the interaction between multiple sensory stimuli related to oral functions and published in English were identified and extracted via three electronic databases and reviewed according to the Preferred Reporting Items for Systematic Reviews and Meta-Analyses. Nine primary studies were eligible to be included in this review. Three studies investigated the interaction of intraoral (i.e., sensorimotor, taste, and noxious) stimuli. Five studies investigated the interaction between intraoral and extraoral (i.e., auditory, olfactory, and visual) stimuli). One study investigated the audio-visual interaction on dental fear. The studies showed great diversity in the experimental design of crossmodal interaction. In terms of the brain features related to the interaction, the somatosensory and motor areas were mostly reported in the studies. The studies imply the potential application between oral neuroscience and oral healthcare, such as prosthodontic treatment and food selection. Still, the findings should be carefully interpreted due to the diversity in crossmodal interaction and inconsistency of experimental design between the studies. The current systematic review revealed that there have not been consistent findings about the underlying brain mechanisms due to the diversity in the experimental design of crossmodal interaction.

## Introduction

Our ‘oral sense’ is fundamentally a multisensory experience. The individual intraoral sensation is formed by a specific sensory pathway. For example, the sense of texture and taste of food is shaped by the tactile and the gustatory pathway via the mechanoreceptors and chemoreceptors, respectively. Furthermore, these primary sensory stimuli are integrated to form a holistic experience of food intake. For example, the somatosensation of texture (i.e., light touch) and pressure (i.e., deep touch) is integrated into oral stereognosis, a somatoperception about the perception of intraoral objects (Haggard & de Boer, 2014). During chewing, the sound of food (e.g., crispy or crunching) may affect our perception of its flavor (Spence, 2015). The viscosity of food, which relates to the kinesthesis of the masticatory system, may be associated with the perception of its taste (Verhagen & Engelen, 2006). Furthermore, the crosstalk between multiple sensory experiences may affect patients’ experience of dental treatment. For example, the sensation of pain, the sound of the drill, and the presence of the needle are all associated with individual dental anxiety (Oosterink, de Jongh, & Aartman, 2008). Until now, the role of multisensory processing in dental practice and oral health has remained unclear.

Multisensory integration refers to ‘the neural integration or combination of information from different sensory modalities. The integration further gives rise to ‘changes in behavior associated with the perception of and reaction to those stimuli’ (Holmes, Calvert, & Spence, 2009). Therefore, multisensory processing of sensory information is associated with the interaction between multiple unimodal processing. Such a crossmodal interaction is different from unimodal processing, as demonstrated by neuroimaging research. For example, when individuals received unimodal auditory or visual stimuli, there was higher brain activation at the auditory and visual cortices, respectively. However, when both stimuli were given in when congruent meanings (e.g., matching between speech and text), there was higher brain activation in the superior temporal gyrus (STG), compared to the brain activation of unimodal stimuli (Calvert, Campbell, & Brammer, 2000). The findings revealed that the STG may participate in a crossmodal (here auditory and visual) interaction or ‘binding’ between these two sensory modalities. At present, neuroimaging studies have revealed that the superior and middle temporal gyri, the thalamus, the insula, and the inferior frontal gyrus may be associated with multisensory integration (Scheliga et al., 2023).

In terms of oral neuroscience, there have been review studies on the brain mechanisms associated with a specific part of oral function, including acute dental pain, chronic orofacial pain, mastication, swallowing, taste, and dental anxiety and fear (Lin, 2021). However, the issue of multisensory integration in oral functions has been largely ignored. While a majority of studies focus on the multisensory processing related to speech (Skipper, Devlin, & Lametti, 2017), the brain mechanisms of non-speech oral multisensory processing have remained unknown. To bridge this gap, the current review aims to investigate the brain mechanisms of non-speech oral multisensory processing, by systematically reviewing the neuroimaging studies on the interaction of multiple sensory modalities in healthy adults. This study focuses on the following three aims:

1. The brain mechanisms associated with oral multisensory processing, as reported by previous neuroimaging studies, will be summarized.
2. The pros and cons of the research methods, especially the experimental paradigms of cross-modal stimulation, will be analyzed.
3. The link between neuroimaging research and translational application in dental practice and oral science will be discussed.

## Methods

### Eligibility criteria

The eligibility criteria of the included studies were defined according to the following features, based on the guideline of the Preferred Reporting Items for Systematic reviews and Meta-Analyses (PRISMA) (Page et al., 2021):

1. Participants: The review included only the studies that investigated human subjects.
2. Intervention/Comparison: The review included both observation research (without an experimental intervention) and experimental research. Specifically, if the study has a treatment (e.g., a drug-induced effect) applied to the participants, only the pre-treatment findings will be focused on in the review. If the study compares a patient group and healthy controls, only the findings of healthy controls will be focused on in the review.
3. Outcomes: The review focused on the brain features identified by the neuroimaging investigation, including magnetic resonance imaging (MRI), magnetoencephalography (MEG), electroencephalogram (EEG), transcranial magnetic stimulation (TMS), and functional near-infrared spectroscopy (fNIRS). Structural (e.g., grey matter volume) or functional (e.g., brain activation during task conditions) features should be reported in the studies. The studies should investigate at least two different unimodal stimuli. The studies need to include (a) at least one of the sensory modalities that are associated with oral sensorimotor processing, such as tactile, taste, and mastication, which relates to proprioception and sensory feedback, or (b) non-oral stimuli that are associated with the behavior of food selection or dental treatment (e.g., dental anxiety and fear). Notably, a typical study of crossmodal interaction may include a ‘bimodal’ condition, i.e., the condition when both unimodal are concurrently applied (Calvert & Thesen, 2004). In this review, either the studies with or without a bimodal condition will be included.

### Search methods

Three electronic databases/indices were searched with the eligibility criteria: PubMed, ProQuest, and Web of Science, by the single author. The following combination of keywords was used for the search: (MRI OR neuroimaging) AND (multisensory OR multi-sensory OR crossmodal* OR cross-modal*) AND (brain OR cortical OR cortex OR cerebell*) AND (tooth OR teeth OR dental OR oral OR orofacial OR intraoral OR mouth OR tongue) (Table 1). No limitation on publication time was applied (until 2024.8.1). Only the articles published in English were included.

**Table 1.** Strategy of keyword search for PubMed (by 2024.8.1)

| Search number | Query | Results |
| --- | --- | --- |
| 1 | MRI OR neuroimaging | 944930 |
| 2 | multisensory OR multi-sensory OR crossmodal* OR cross-modal* | 13864 |
| 3 | brain OR cortical OR cortex OR cerebell* | 2711751 |
| 4 | tooth OR teeth OR dental OR oral OR orofacial OR intraoral OR mouth OR tongue | 1799807 |
| 5 | #1 AND #2 AND #3 AND #4 | 70 |

### Data collection

The following seven items of study features were extracted from the full text of the included articles by the single author: (1) the source of the study, including the name of the first author and the year of publication, (2) the number of participants, including the number for each subgroup based on the study design, (3) the range or mean±standard deviation of the participants’ age, (4) the method of neuroimaging, (5) the condition of unimodal sensory stimuli, (6) the condition of bimodal stimuli, and (7) the approach to analyze crossmodal interaction. If the information for a data item was labeled ‘not available’ (n.a.) if it cannot be identified in an article.

### Data synthesis

Most of the current neuroimaging studies focused on the spatial pattern of brain activation; therefore, the review investigated the brain regions associated with crossmodal interaction, as the primary outcome. The brain regions reported in individual studies were first identified from the tables and figures of each study. Subsequently, the findings from all the studies were summarized by tabulation. The hemisphere (left, right or bilateral) of the brain regions was identified. Notably, because the included studies diverged greatly in the research variables (i.e., sensory modalities) and study design, no further meta-analysis was performed due to the great between-study heterogeneity.

### Study risk of bias assessment

The assessment of the risk of bias (RoB) was based on the design of the Newcastle-Ottawa scale (NOS) for nonrandomized studies (Wells et al., 2000). The original items were customized for evaluating the RoB of neuroimaging studies, including the following five items: (1) the systemic condition and (2) the oral condition of participants, which were related to the bias of selection and representativeness, (3) the design of task comparison (e.g., if a proper control condition was used), which was related to the bias of comparability, and (4) the correction of head motion and (5) thresholding of imaging results, which were related to the bias of outcome. Each item was assessed as high or low risk, or uncertain.

## Results

### Study selection

Based on the search of three electronic databases/indices, 138 records were identified and 104 records were further screened for their eligibility. In total, 95 of them were excluded (see Supplementary Material for the reasons for exclusion). Nine studies were eligible to be included in this review (Figure 1). The studies were categorized into three groups according to the modalities they investigated (Table 2):

**Figure 1.**
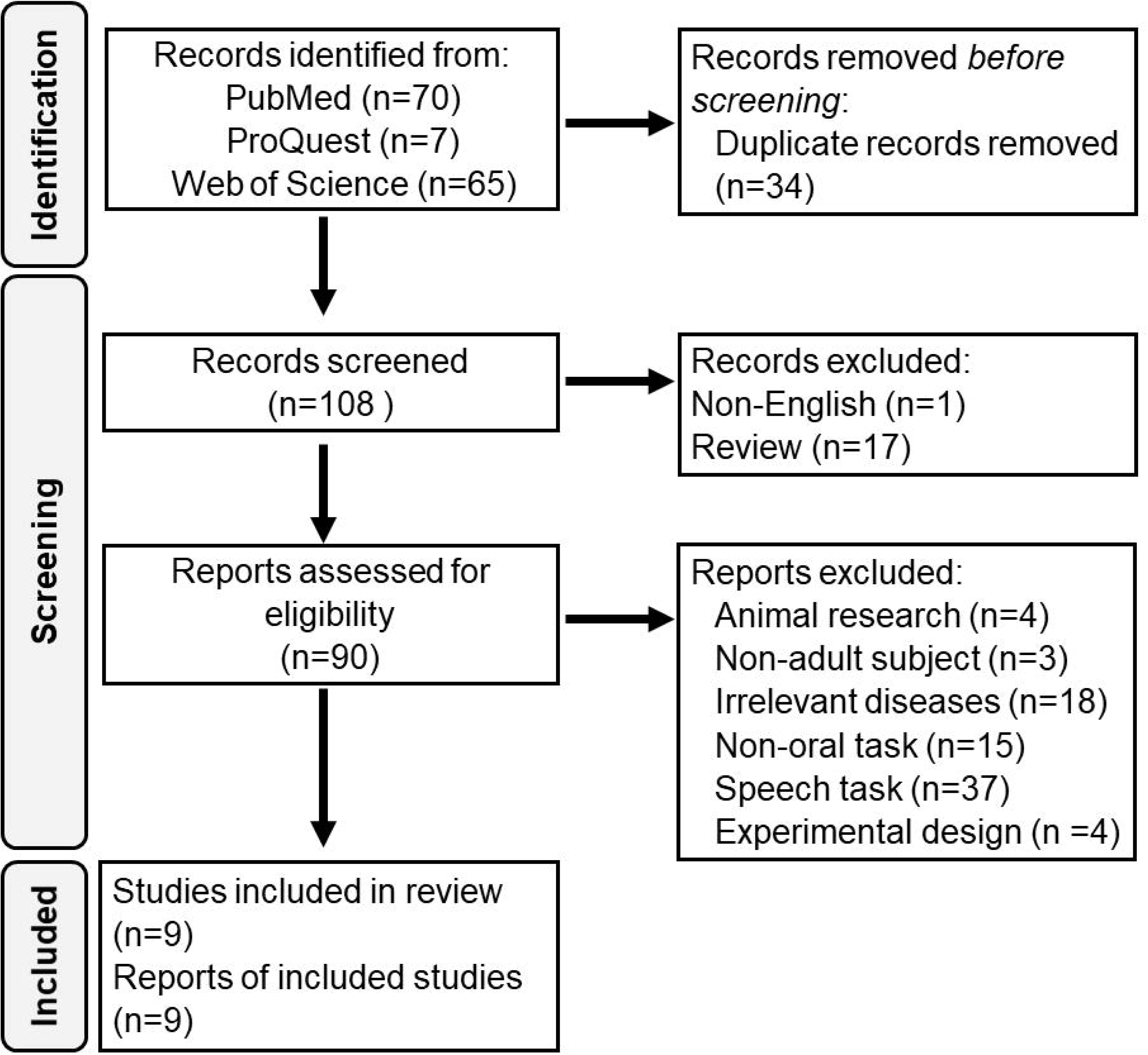
Flow diagram of searching and screening of eligible articles.

**Table 2.**
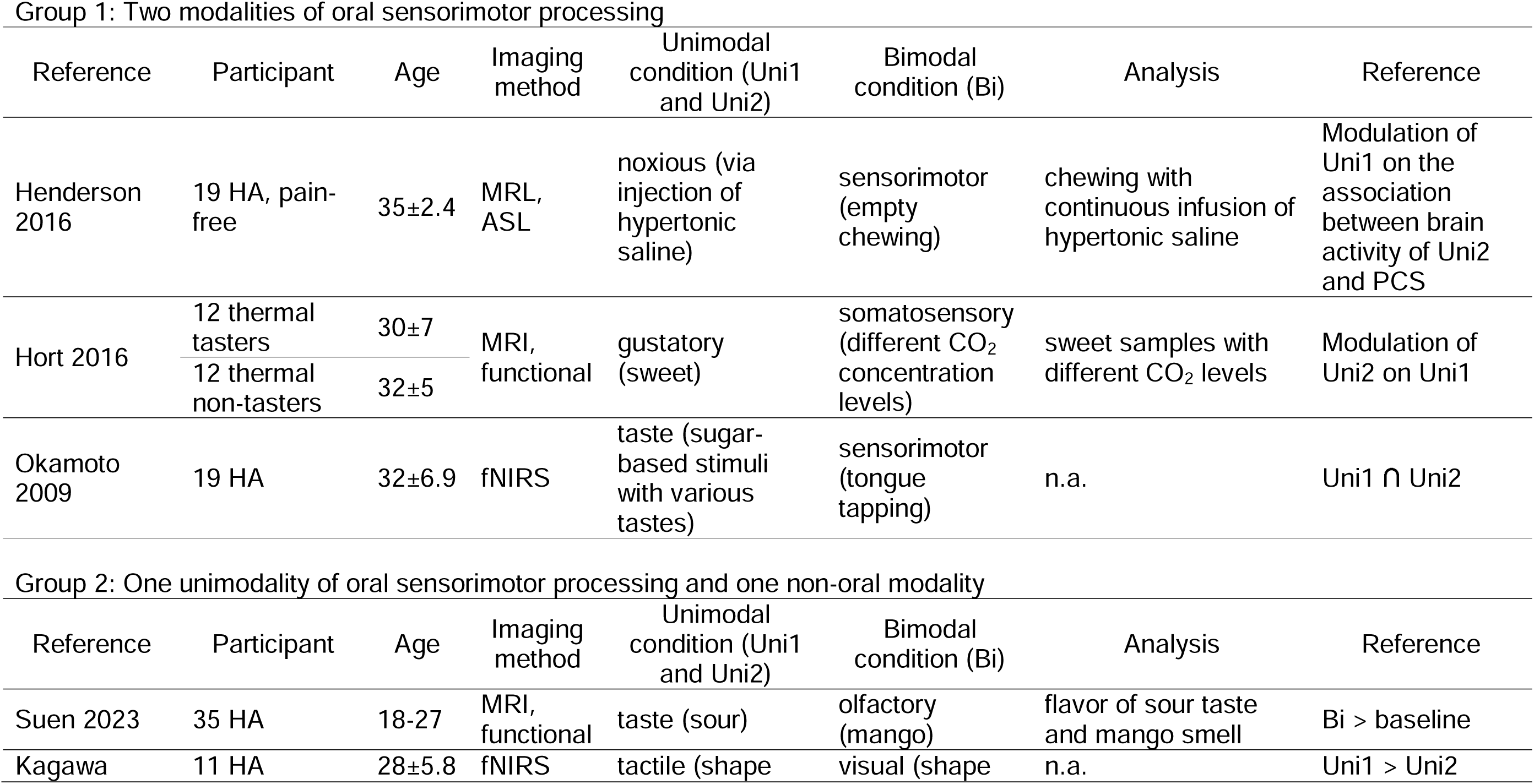
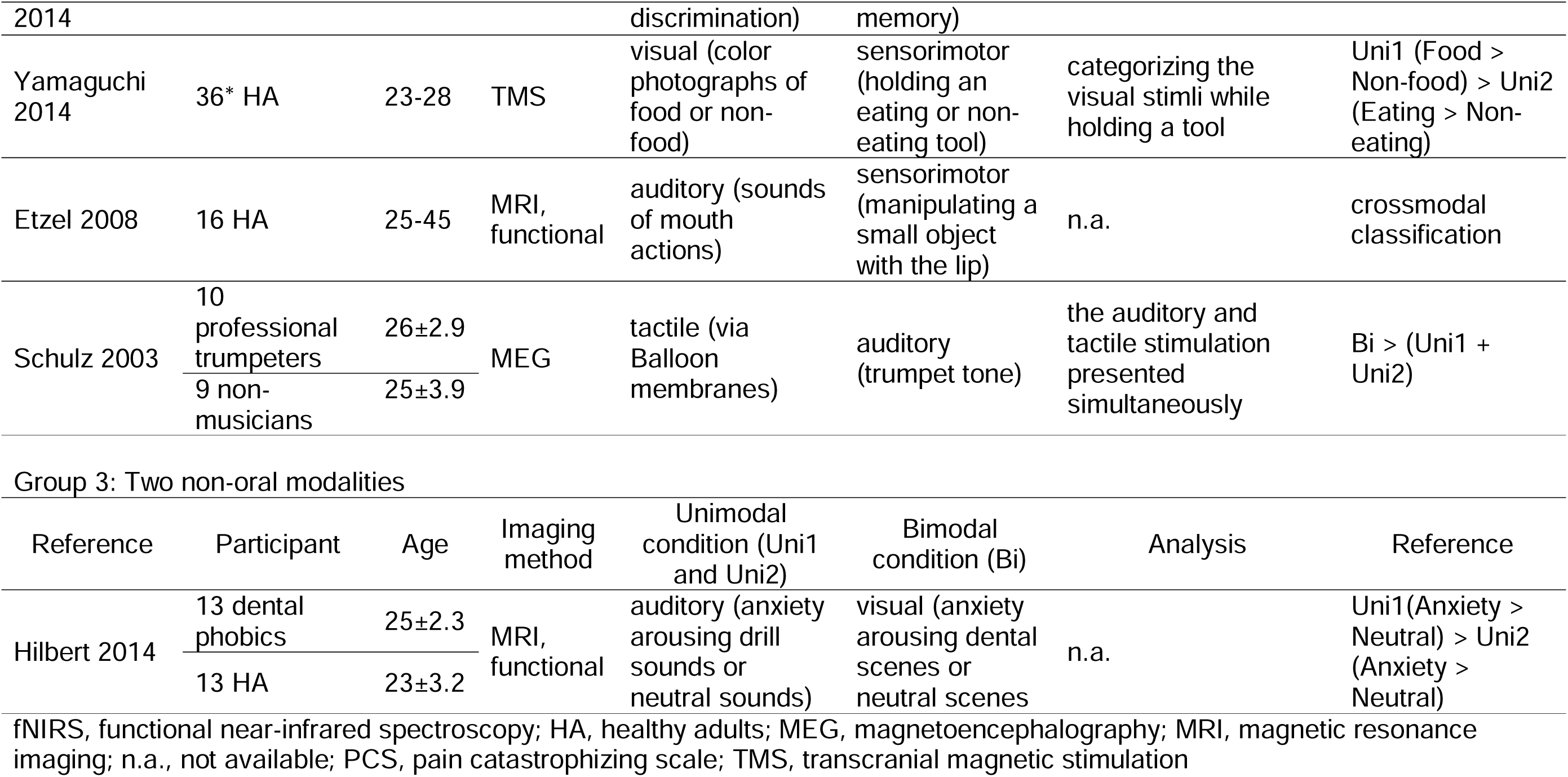
Study characteristics of the included articles.

1. The first group of studies focused on the interaction between two modalities of oral sensorimotor processing, including somatosensation and taste (Hort, Ford, Eldeghaidy, & Francis, 2016), nociception and jaw movement (Henderson et al., 2016), taste and tongue movement (Okamoto, Dan, Clowney, Yamaguchi, & Dan, 2009).
2. The second group focused on the interaction between one modality of oral sensorimotor processing and another non-oral sensory modality, including taste and olfaction (Suen et al., 2021), vision and somatosensation (Kagawa et al., 2014; Yamaguchi, Nakamura, Oga, & Nakajima, 2014), somatosensation and audition (Schulz, Ross, & Pantev, 2003), and audition and mouth actions (Etzel, Gazzola, & Keysers, 2008).
3. The third group focused on the interaction between two non-oral sensory modalities, i.e., vision and audition, which was associated with the experience of dental anxiety and fear (Hilbert, Evens, Maslowski, Wittchen, & Lueken, 2014).

Some studies, though including task conditions of oral sensorimotor processing, were excluded due to the lack of analysis on crossmodal interaction. For example, a recent study on the association between orofacial pain and visual processing was excluded from this review because only one modality of sensory stimuli (i.e., visual) was investigated (Schulz et al., 2003). Another study on stereognosis (Schumann-Werner et al., 2023) was excluded because although the study included two tasks of sensory stimuli (oral and manual assessments of stereognosis), it focused on the common mechanisms shared by both (oral vs. manual) assessments, rather than the interaction of sensory processing of oral stereognosis.

### Study characteristics

In terms of the participants, three studies included an investigation of different groups of participants, including thermal tasters vs. non-thermal tasters (Hort et al., 2016), patients with dental phobic vs. healthy controls (Hilbert et al., 2014), and professional musicians vs. non-musicians (Schulz et al., 2003). All the studies investigated adults only (Table 2). In terms of the sensory modalities being investigated, sensorimotor processing associated with kinesthesia and proprioception, including jaw movement (Henderson et al., 2016), tongue movement (Okamoto et al., 2009), holding eating tools (Yamaguchi et al., 2014), and lip movement (Etzel et al., 2008), was the most frequently investigated (four times) modality. Gustation (Hort et al., 2016; Okamoto et al., 2009; Suen et al., 2021), vision (Hilbert et al., 2014; Kagawa et al., 2014; Yamaguchi et al., 2014), audition (Etzel et al., 2008; Hilbert et al., 2014; Schulz et al., 2003), and somatosensation (Hort et al., 2016), including tactile stimulation (Kagawa et al., 2014; Schulz et al., 2003), were investigated three times, and nociception and olfaction were investigated once, respectively (Table 2). Among the nine studies, five of them used magnetic resonance imaging (MRI) to study the brain features, including four function MRI studies and a study of arterial spin labeling (ASL). Two studies applied fNIRS. Finally, TMS and MEG were applied in one study, respectively (Table 2). In terms of the analyses of crossmodal interaction, the nine studies showed a great divergence. The brain activation of bimodal condition was investigated in two studies (Schulz et al., 2003; Suen et al., 2021). Interaction between two unimodal conditions was investigated in two studies (Hilbert et al., 2014; Yamaguchi et al., 2014).

### Risk of bias in studies

The results of the RoB analysis were summarized in Table 3. In general, all the studies showed a low to moderate degree of RoB, judging from the five items of the RoB assessment (Table 3). Among the items, most of the studies have conducted correction of head motion and applied a proper baseline condition for experimental control. While most of the studies have explicitly stated the inclusion and exclusion criteria of the systemic conditions of participants, very few studies have investigated the oral conditions of the participants. Activation of the premotor cortex (PMC) was noted in one study on lip movement (Etzel et al., 2008).

**Table 3.**
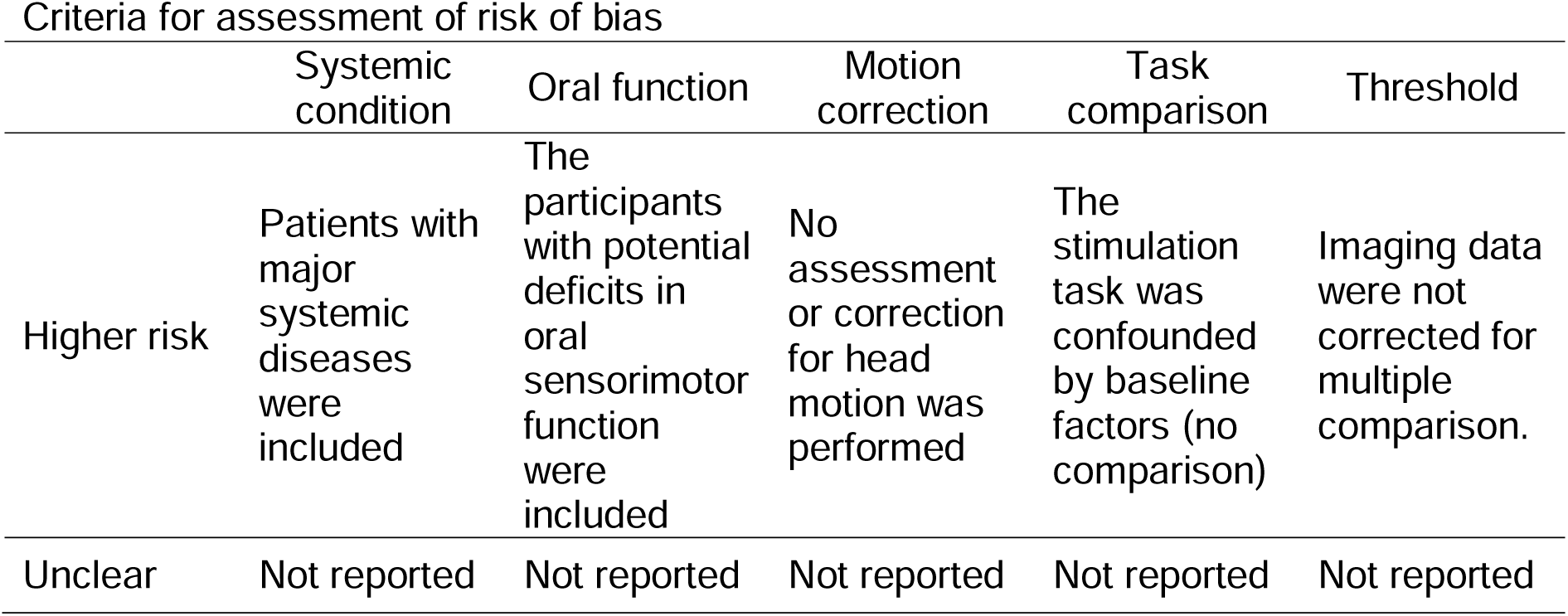
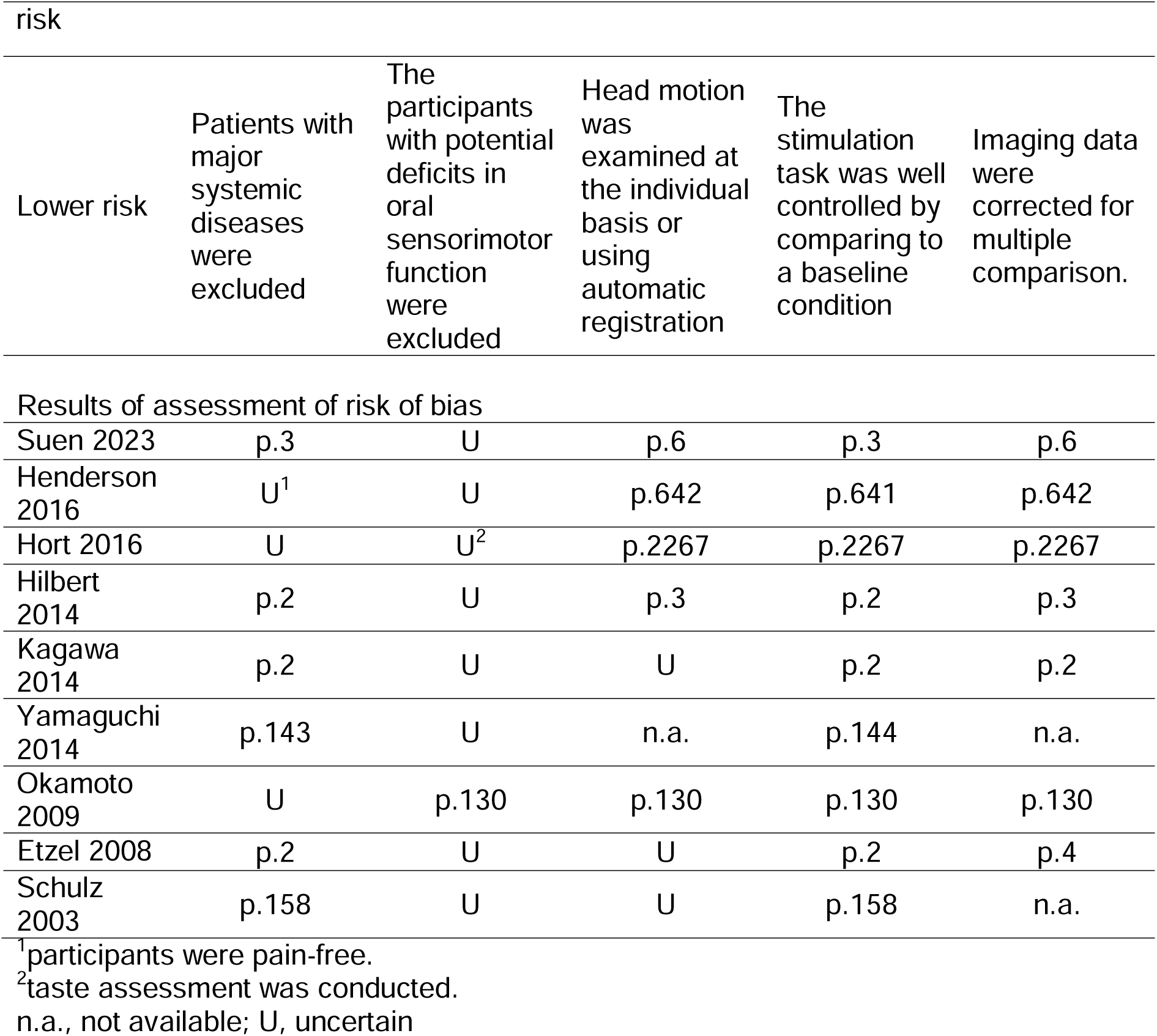
Results of risk of bias assessment.

### Results of individual studies

As shown in Table 2, all the studies have reported statistically significant findings for the cross-modal interaction between the sensory modalities. The findings of brain features were further presented in Table 4. The following findings were identified and tabulated: Table 2 from (Suen et al., 2021), Table 4 from (Hort et al., 2016), Table 2 from (Etzel et al., 2008), Figure 6 from (Kagawa et al., 2014), Figure 3 from (Henderson et al., 2016), Figure 2 from (Okamoto et al., 2009), Figure 2 from (Schulz et al., 2003), and p.144 from (Yamaguchi et al., 2014). Each of the studies had different focuses on the sensory modalities and there existed a great heterogeneity in the approach of analyzing crossmodal interaction.

**Figure 2.**
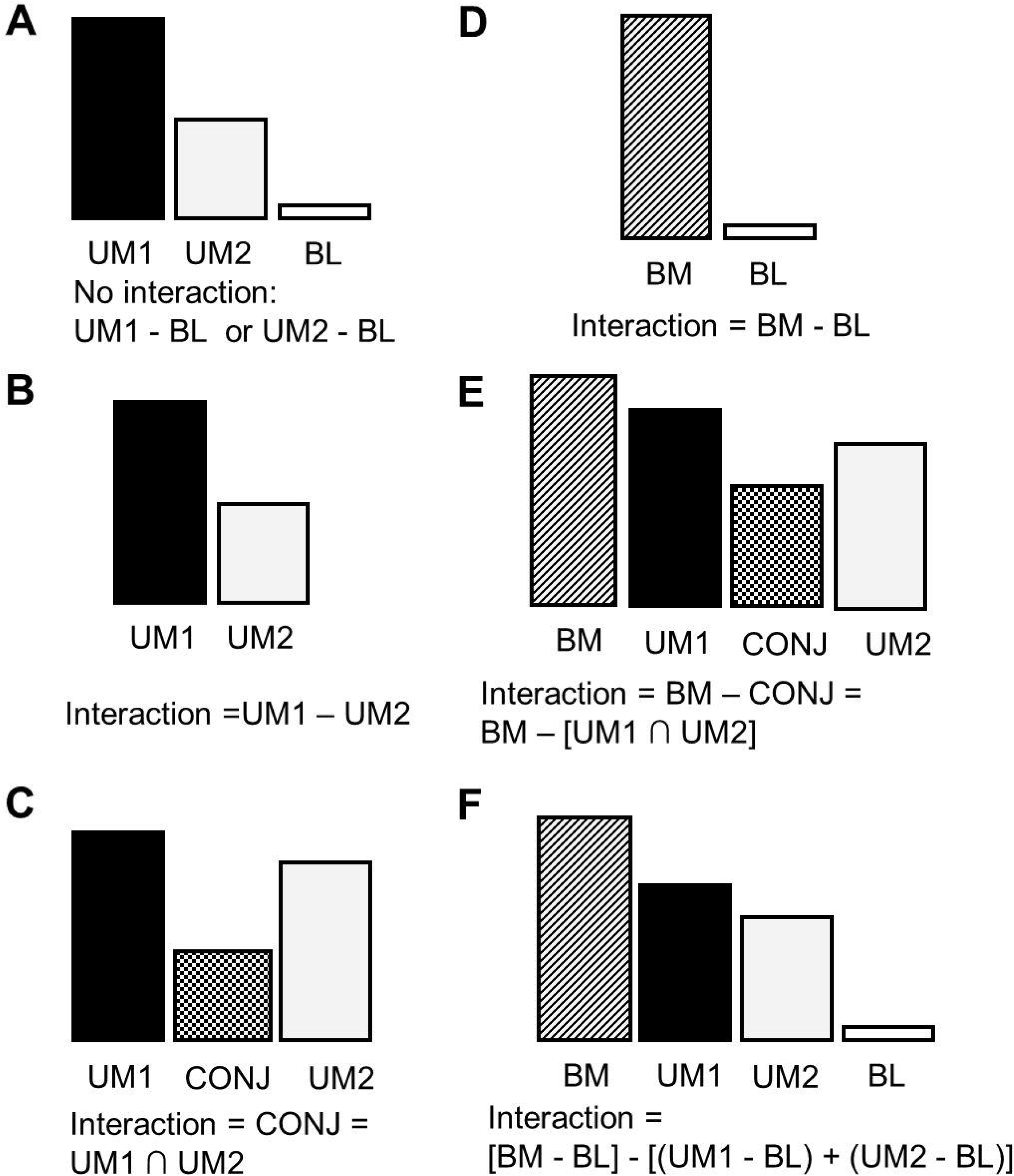
Study design of crossmodal interaction. (A) The design investigates the brain features associated with each of the unimodal stimuli (UM1 and UM2) by contrasting with the baseline (BL) condition, however, not showing the interaction between the two stimuli. (B-C) The designs investigate either the differential pattern of activation between the two modalities (B) or the common pattern between the two modalities (C). (D-F) The designs include a bimodal condition (BM), which is contrasted to the baseline (D) or the conjunction of the unimodal condition (E). The interaction can be contrasted by detecting the change relative to the baseline condition, respectively for bimodal and each unimodal condition.

**Table 4.** Brain regions associated with crossmodal interaction.

Group 1: Two modalities of oral sensorimotor processing
| Study | Modalities |  | M1 | PMC | Basal ganglia | Cerebellum | S1 | S2/ RO | PPC | Olfactory | Visual | Insula | CC | PFC |
| --- | --- | --- | --- | --- | --- | --- | --- | --- | --- | --- | --- | --- | --- | --- |
| Henderson | noxious | sensori-motor | L |  |  | R |  |  |  |  | R |  |  | R |
| Hort 2016 | gustatory | somato-sensory | R |  |  |  | B | B |  |  |  |  | B |  |
| Okamoto 2009 | gustatory | sensori-motor | B |  |  |  | B |  |  |  |  |  |  |  |

Group 2: One unimodality of oral sensorimotor processing and one non-oral modality
| Study | Modalities |  | M1 | PMC | Basal ganglia | Cerebellum | S1 | S2/ RO | PPC | Olfactory | Visual | Insula | CC | PFC |
| --- | --- | --- | --- | --- | --- | --- | --- | --- | --- | --- | --- | --- | --- | --- |
| Suen 2023 | gustatory | olfactory | R |  | B | B | L | R |  | L |  | R | L | R |
| Kagawa 2014 | tactile | visual |  |  |  |  |  |  | L |  | B |  |  |  |
| Yamaguchi 2014 | visual | sensori-motor | L |  |  |  |  |  |  |  |  |  |  |  |
| Etzel 2008 | auditory | sensori-motor |  | B |  |  |  |  |  |  |  |  |  |  |
| Schulz 2003 | tactile | auditory |  |  |  |  | B |  |  |  |  |  |  |  |

Group 3: Two non-oral modalities<sup>1</sup>
| Study | Modalities |  | M1 | PMC | Basal ganglia | Cerebellum | S1 | S2/ RO | PPC | Olfactory | Visual | Insula | CC | PFC |
| --- | --- | --- | --- | --- | --- | --- | --- | --- | --- | --- | --- | --- | --- | --- |

As shown in Table 4, the primary motor cortex (M1) and the primary somatosensory cortex (S1) were most frequently reported in the studies. Notably, activation of these regions was associated with touch or oral sensorimotor processing, including jaw movement (Henderson et al., 2016), tongue movement (Okamoto et al., 2009), holding eating tools (Yamaguchi et al., 2014), and somatosensory stimuli (Hort et al., 2016; Suen et al., 2021). In contrast, the brain regions associated with crossmodal integration, including the STG, the insula, and the prefrontal cortex (PFC), were seldom reported in the studies (Table 4).

## Discussion

### Brain mechanisms associated with non-speech oral multisensory processing

A recent neuroimaging meta-analysis of 49 studies revealed that the processing of multi-modal (i.e., auditory, visual, tactile, gustatory, and olfactory) sensory stimuli was associated with brain activation in the temporal lobe, the thalamus, the right insula, and left inferior frontal cortex (Scheliga et al., 2023). The findings are not consistent with the pattern of brain activation reported in the current review (Table 4). Here, a more diverse pattern of brain activation was found across the studies. The only brain regions that were repeatedly identified across the studies were the somatosensory area, including S1 and the secondary somatosensory cortex, which were identified in four studies, and the motor area (including M1, PMC, the basal ganglia, and the cerebellum), which were found in five studies. In contrast to the previous meta-analysis (Scheliga et al., 2023), there was few ‘multisensory region’ (e.g., the temporal lobe) identified. Only one study reported the posterior parietal cortex (PPC) (Kagawa et al., 2014), which was identified for multisensory integration in previous research (Sereno & Huang, 2014). In addition, the brain regions associated with associative learning and attentional control, e.g., the PFC, the insula, the thalamus, and the insula, which were also associated with multisensory integration (Chen et al., 2015; Zikopoulos & Barbas, 2007), were not consistently found here (Table 4).

Nevertheless, the current findings disclose a critical pattern that the sensorimotor area may play a key role in oral multisensory processing. This is partly because in most of the included studies, a task condition of oral sensorimotor processing, such as passively perceiving the taste of food or actively moving jaw or tongue, was involved. Notably, the ‘unimodal’ stimulus in the mouth may not induce only a single sensory pathway. For example, when individuals tried to perceive the taste of food, tongue movement helps the contact between tastants and chemical receptors on the tongue. Therefore, in all the studies including taste stimuli, there was activation in the sensorimotor area (Hort et al., 2016; Okamoto et al., 2009; Suen et al., 2021). The findings echoed the concept that sensorimotor processing plays a key role in shaping the ‘mouth experience’, i.e., individuals actively explore the intraoral conditions by integrating sensory feedback and motor activity (Haggard & de Boer, 2014).

### Pros and cons of experimental design of crossmodal interaction

The review showed great heterogeneity in the study design (Table 2). All nine studies investigated different combinations of sensory modalities. For the same sensory modality, different task conditions were used. For example, jaw, tongue, and lip movement may induce different degree of kinesthesia and proprioception (Table 2). Furthermore, the studies used different approaches to analyze crossmodal interaction, and the divergence of the analytic approach greatly affects how brain features are interpreted. The inconsistency in experimental design may explain the absence of brain activation of primary sensory modality. For example, the study by Okamoto et al. analyzed the conjunction of activation between taste and sensorimotor tasks (Okamoto et al., 2009). Because the intersected regions were identified, the brain activation specific to one task (e.g., taste) but not to the other (e.g., tongue tapping) was not shown in the conjunction. The study by Hort et al. investigated the modulational effect of somatosensory stimuli on gustation with three experimental conditions (no, low, and high concentration of CO_2_) (Hort et al., 2016). Critically, gustatory stimuli were delivered in all three conditions, and therefore, a contrast between these conditions may not reveal brain activation specific to gustation.

In general, to investigate multisensory integration does not mean merely including multiple tasks of individual sensory stimulation. The key to ‘integration’ is the interaction between these multi-modal stimuli. As shown in Figure 2A, many neuroimaging studies investigated the effect of two unimodal stimuli, and each of them was contrasted with baseline (control) conditions. However, the findings only revealed the brain features related to each of the unimodal stimuli. The design of Figure 2B investigated the difference between two unimodal stimuli. The approach may only reveal the brain processing involved in one sensory pathway but not in another. In contrast, the design of Figure 2C better reflects the common pattern of brain activation by identifying the conjunction of brain regions reported in individual unimodal conditions (Calvert & Thesen, 2004). The crossmodal interaction may be better contrasted by including a condition of ‘bimodal’ stimulation, i.e., two primary sensory modalities were delivered concurrently. The simplest design is to contrast the brain activation of a bimodal condition to a baseline condition (Figure 2D). However, the results may only reflect the change of baseline mental activity, such as heightened attention, which is common to both unimodal stimuli and not relevant to crossmodal interaction. To better capture such an interaction, one may investigate the difference between the bimodal condition and the conjunction between two unimodal conditions (Figure 2E) and identify the effect of interaction using a two-factor design (Figure 2F). The latter design would better reflect the ‘interaction’ in a statistical sense, i.e., the change of one effect (from the first sensory stimuli) associated with another effect (from the second sensory stimuli) (Calvert & Thesen, 2004).

### Clinical implications

The current review showed that all the studies only focused on healthy adults and did not investigate oral multisensory processing related to oral diseases (Table 2). However, the findings disclosed that even in healthy adults, the shaping of multisensory experience related to oral functions is very complicated – it is associated not only with the peripheral structure of the stomatognathic system but also the central nervous system. The findings may have three major impact on the clinical practice of oral medicine:

1. At present, most of the assessment of oral function focused on a specific sensory or motor activity. As demonstrated in this review, many of these unimodal stimuli interacted with each other and an assessment of the multisensory experience of oral function should be considered.
2. Multisensory processing is associated with brain mechanisms, and therefore, individual differences in brain features would play a key role in their multisensory experience of oral functions. This point is especially important to elderly people who demonstrated age-related changes in brain function and structure. Moreover, the findings imply that altered oral multisensory experience may occur in patients with neurological deficits, e.g., neurodegenerative disorders.
3. Furthermore, recent neuroimaging and animal findings have highlighted the plasticity of the brain, which relates to the change in oral function (Sessle, 2019; Trulsson et al., 2012). It is noteworthy that multisensory experience can be sculpted by training (Johansson, 2011; Paraskevopoulos & Herholz, 2013). Therefore, in addition to the improvement of a single oral function (e.g., masticatory performance or maximal biting force), the improvement of crossmodal processing of sensory information should be a critical goal.

### Limitations and further considerations

The findings from this review should be interpreted cautiously with the following limitations. First, the divergent pattern of brain activation associated with oral multisensory processing, as shown in Table 4, resulted from qualitative rather than quantitative analysis (e.g., the coordinate-based meta-analysis) of neuroimaging findings. Because there exists a great heterogeneity in study design (Table 2), no further statistical approach was used to synthesize these patterns of brain activation. Second, as stated in the previous section, there has been no standardized approach to the design of the task conditions for crossmodal interaction. Some approaches may better reflect the interaction than others (Calvert & Thesen, 2004). Therefore, the brain features revealed by a study should be carefully interpreted according to the study design. Third, most of the studies included here were not designed for assessing clinical symptoms or treatment effects. Therefore, the findings should not be overdrawn for clinical assessment or diagnosis.

## Conclusions

Though neuroimaging methods have been applied to investigate the brain features of non-speech oral multisensory processing, there have not been consistent findings about the underlying brain mechanisms due to the diversity in the experimental design of crossmodal interaction.

## Supporting information

Supplementary_Multisensory_oral_exclusion_20230628

Supplementary_Multisensory_oral_PRISMA_2020_checklist_20230627

## Author contributions

C-S Lin contributed to the conception, design, and data collection of the study, and drafted and finalized the manuscript.

## Funding

C-S Lin is funded by the Ministry of Science and Technology of Taiwan (110-2314-B-A49A-518-MY3).

## Conflict of interest

The author has no conflicts to report.

## Supplementary material

The Supplementary Material for this article can be found online.

## Data Availability

All data produced in the present work are contained in the manuscript

## Notes

### Competing Interest Statement

The authors have declared no competing interest.

